# Evaluation of the Effects of Oxidative Stress and Biomarkers of Inflammation in Cardiomyopathy Sufferers

**DOI:** 10.1101/2024.01.03.23300634

**Authors:** Shivam Tiwari, Om Shankar, Royana Singh, Ajay Kumar Yadav, Anil Kumar Maurya, Umesh Choudhary

**Author notes:** **Correspondence**: Dr. Umesh Choudhary, Department of Anatomy, Institute of Medical Sciences, Banaras Hindu University, Varanasi, India.

## Abstract

**Introduction:** Cardiovascular disease can develop and worsen as a result of inflammation and oxidative stress. The current research looked into the relationship between oxidative damage and biomarkers of inflammation in individuals with cardiomyopathy.

**Methods:** Particular kits for ELISA were used to measure the serum concentrations of CRP, TNF-α, IL-6, and NT-proBNP. These specific ELISA kits are based on sandwich enzyme immunoassay techniques whose results are quantitative. The accuracy of the tests was established by comparing them to control sera that were included in the kits and had known quantities of the analytes.

**Results:** When compared to individuals without cardiomyopathy (control group), we found that cardiomyopathy patients had significantly higher blood C-reactive protein concentrations (P =<0.0001). When compared between control case and cardiomyopathy patients, then find that cardiomyopathy patients had significantly higher concentrations of tumour necrosis factor-alpha (TNF-alpha) (P =<0.0001). TNF-α associated favourably with malondialdehyde (P =<0.0001, r =0.4524) and glutathione peroxidase (GPX) (P =<0.0001, r =0.8311) in Cardiomyopathy patients. Interleukin-6 was not significantly linked with GPX (P =0.0001, r =-0.1194) in cardiomyopathy patients. In those with cardiomyopathy, there was a strong association (P=<0.0001, r=0.4826) between malondialdehyde and NT-proBNP. Furthermore, we observed that the activity of glutathione peroxidase (GPX) exhibited a significant connection with NT-proBNP (P =<0.0001, r = 0.6084) in all cardiomyopathy patients.

**Conclusions:** In cardiomyopathy patients, but not in normal cases, there is a correlation between inflammatory and oxidative stress indicators. These findings imply intricate cross-talk between the two cellular processes in late-stage cardiomyopathy.

## INTRODUCTION

More and more evidence points to the pathogenesis and development of congestive heart failure (CHF) as being influenced by inflammatory processes.^1,2^ Myocardial dysfunction brought on by inflammation can result in endothelial dysfunction and cardiac cachexia. Blood leukocytes, platelets, endothelial cells, failing myocardial cells, the liver, and the lungs all emit inflammatory markers.^1,3^

Patients with heart failure have been found to have higher levels of inflammatory indicators (C-reactive protein (CRP), cytokines) and white blood cell count (WBC) with immune system activation markers.^4,7^ As mitral valve disease (MMVD) progresses, there is a reduction in the levels of cytokines such as IL-2, IL-7, and IL-8.^14^ It has been demonstrated via clinical and experimental studies that oxidative stress plays a major role in the myocardium hypertrophy and heart failure.^1,17^

We hypothesised that in situations involving cardiomyopathy, which oxidative and intolerant stress labels have been linked either with DCM or other type of cardiomyopathy. We assessed designated seditious and oxidative stress markers to put the theory to the test. To put the theory to the test, we measured designated seditious and oxidative stress labels.

## METHODS

Two hundred and Two cases were examined in the current study. There was no indication of allergic illnesses, cancer, renal disease, endocrine problems, or infectious diseases, thus one hundred and one cases were included as a healthy control group. With one hundred and one case healthy control cases and one hundred and one patients with either dilated cardiomyopathy or another kind of cardiomyopathy were prospectively recruited.

According to their histories, physical examinations, regular laboratory findings (hematology, biochemical profiles), and NT-proBNP levels, the one hundred and one patients cases were deemed as a cardiomyopathy case. Under the guidance of Dr. Om Shankar, Department of Cardiology, IMS, BHU, DCM, and other cardiomyopathy, heart failure were identified. Patients were divided into groups according to whether they had congestive cardiomyopathy, other cardiomyopathies. Overall the patient group consisted of a total of 101 patients diagnosed with cardiomyopathy. The control group consisted of 101 patients without cardiomyopathy.

The patient with cardiomyopathy granted their signed consent. All methods were authorised through the Institute’s Ethical Committee. Ethical approval for “Genetic basis of cardiomyopathy in eastern Uttar Pradesh population” was obtained vide letter number Dean/2022/EC/3824 dated 15.04.2023. Blood samples from one hundred and one patients visited to the department of Cardiology of Hospital due to the severity of symptoms were obtained, carried to the Multidisciplinary Research Unit of Institute stored at −80 degrees centigrade, and processed under biosafety-compliant procedures by appropriately trained personnel. Medical histories were obtained from hospital records. Samples from one hundred and one apparently healthy patients with no cardiac issue within the last month were included as the control group.

### Laboratory Work

All laboratory analyses required blood samples, which were taken from the case as well as control. Serum tubes (Vaccuette; Hindustan Syringes & Medical Devices LTD., Ballabgarh, Faridabad, India) were centrifuged for 12 minutes at 1300 g after being left at room temperature for 30 minutes. Once the trial was over, the blood used to measure the levels of CRP, TNF-α, and IL-6 was frozen at −80 °C and analysed in batches. On the day of collection, serum biochemical parameters were measured. Samples were collected in EDTA tubes (Vacuette; Hindustan Syringes & Medical Devices LTD., Ballabgarh, Faridabad, India) to determine plasma concentrations of NT-proBNP and MDA. The plasma samples obtained from these tubes were immediately frozen at −80 °C until analysis and then centrifuged for 15 min. at 1500 g at 4 °C. At the conclusion of the investigation, batch measurementsof NT-proBNP and MDA concentrations were taken. To evaluate the activity of whole blood glutathione peroxidase (GPX), blood samples were collected into tubes containing the anticoagulant lithium heparin (Vacuette; Hindustan Syringes & Medical Devices LTD., Ballabgarh, Faridabad, India). Following the heparinization of the entire blood, aliquots were prepared and promptly frozen at 80 °C for batch analysis.

### Measurement of CRP, TNF-α, and IL-6 concentrations

Commercially available, specific ELISA kits (CRP ELISA; Cat No. ELK1040, ELK Biotechnology Co.,LTD. Hubei, P.R.C.; TNF-α alpha Cat No. ELK1190, ELK Biotechnology Co.,LTD. Hubei, P.R.C.; and IL-6 Cat No. ELK1156, ELK Biotechnology Co.) were used to measure the quantities of blood CRP, TNF-α, and IL-6. The test ranges for TNF-α, IL-6, and CRP were respectively 15.63-1000 pg/mL (sensitivity: 6.5 pg/mL), 7.82-500 pg/mL (sensitivity: 3.2 pg/mL), and 62.4-4000 pg/mL (sensitivity: 26.1 pg/mL). The tests’ accuracy was verified using control sera that were provided in the kits and had known analyte concentrations. TNF-α, IL-6, and CRP had coefficients of variation of 8.9%, 2.75, and 9.8%, in that order.

### Determination of GPX activity

In the experiment, 0.2 mL of serum were utilised. In order to produce a persistent colour, the GSH was forced to react with 5-dithiobis (2-nitrobenzoic acid) [DTNB], which interacts with sulfhydryl groups. GSH content was reported as mol/gHb, and absorbance was measured at 412 nm.

### Determination of MDA and NT-proBNP concentrations

Serum malondialdehyde was extracted and identified as a conjugation with TBA. TCA and centrifugation were used to remove the serum proteins. At 534 nm, the MDA-TBA complex was detected (8).All the three reagent:-1st Reagent (Trichloroacetic Acid 17.5%): To aid in evaporation, 17.5 grammes of TCA were dissolved in 100 ml of distilled water,Reagent 2: (70 percent trichloroacetic acid) and Reagent No. 3 (Thiobarbituric Acid, 0.6%): To prevent evaporation, the reagent was placed in a sealed glass bottle with 600 mg of TBA and 100 ml of distilled water. The reaction was conducted in an 18 x 150 mm Pyrex test tube marked “test & blank,” into which the reagents are taken in equal amount with sample. The tubes were filled with an equal amount of each reagent and sample, mixed thoroughly, and incubated in a boiling bath for 15 twinkles. The tubes were then allowed to cool and stood at room temperature for 20 twinkles. The tubes were also centrifuged for 15 twinkles at 2000 rpm, and the supernatant subcaste was read at 534 nm.The following formula was used to determine the MDA concentration (nmol/ml): Test concentration: 1.56 x 1000000 / Abs (test) - Abs (blank).The NTproBNP enzyme-linked immunosorbent test (ELISA; Cat No. ELK1212, ELK Biotechnology Co.,LTD. Hubei,P.R.C.;) was used to assess plasma NT-proBNP concentrations. The test ranges for NT-proBNP were 39.07-2500 pg/mL and sensitivity is 15 pg/mL. The assays’ precision was verified using control sera, which were added to the kits and had known amounts of the analytes. The coefficients of variance for NT-proBNP were 8.9%.

## RESULTS

In this prospective study, 101 healthy cases served as the control group along with 101 cases of cardiomyopathy (including 43 cases of dilated cardiomyopathy, or DCM, and 58 cases of other cardiomyopathies such hypertrophic cardiomyopathy etc.). When compared to patients without cardiomyopathy (control group), patients with cardiomyopathy had significantly higher blood concentrations of C-reactive protein (P =0.0001), as seen in Table 1. Tumour necrosis factor-alpha (TNF-alpha) concentrations were substantially greater in patients with cardiomyopathy (P =<0.0001) when compared to the control group. In individuals with cardiomyopathy, TNF-α showed a favourable correlation with glutathione peroxidase (GPX) (P =<0.0001, r =0.4524) and malondialdehyde (P =<0.0001, r =0.8311). Interleukin-6 and GPX did not, however, significantly correlate (P =<0.0001, r= −0.1194). Additionally, in those who have cardiomyopathy, NT-proBNP Furthermore, among those with cardiomyopathy, NT-proBNP was associated with malondialdehyde (P=<0.0001, r=0.4826). Also, as shown in Table 2Figure 2, GPX activity was shown to be positively related to NT-proBNP in all cardiomyopathy patients (P =<0.0001, r = 0.6084).

**Figure 1.**
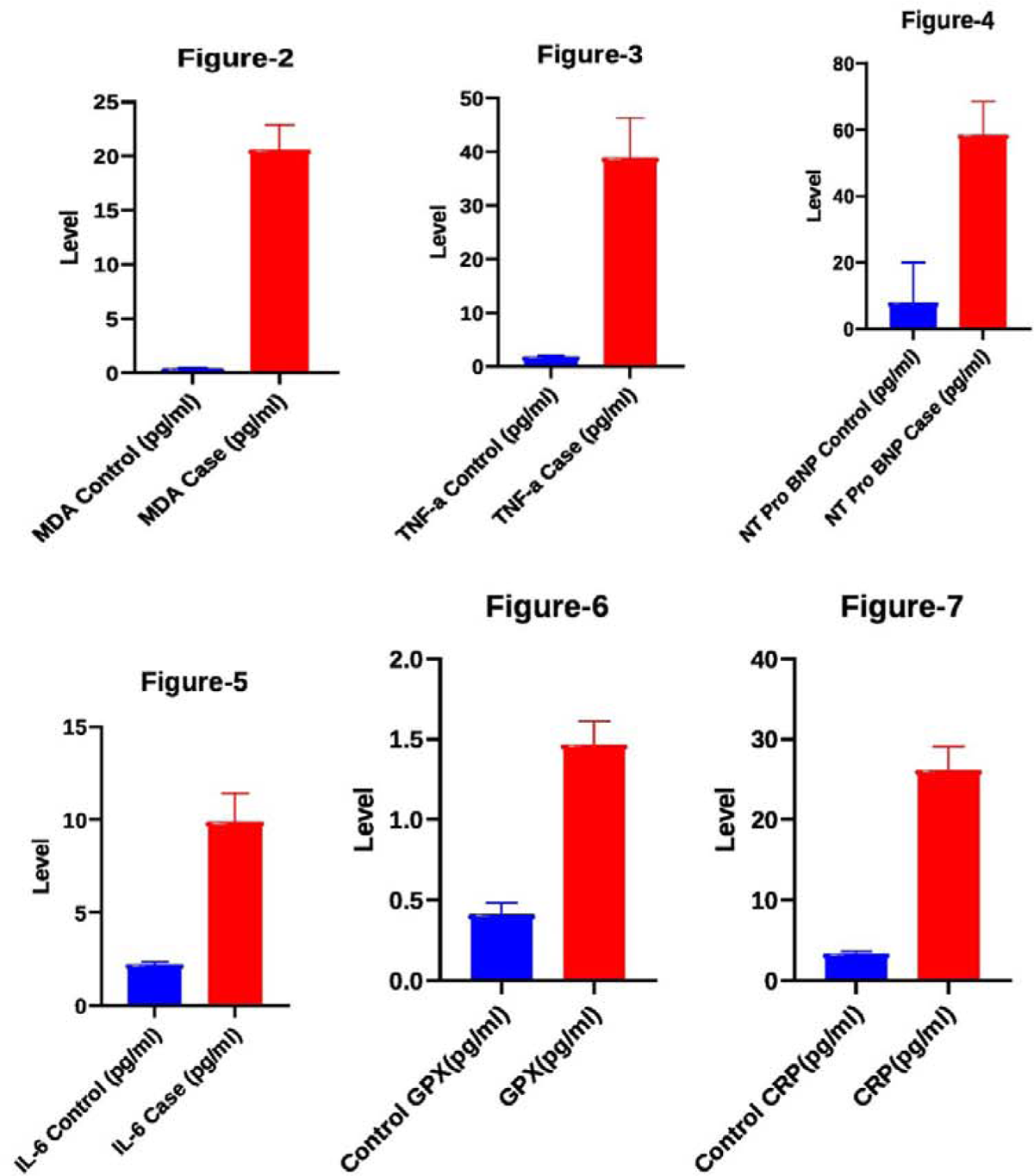
(Figure 2,3,4,5,6) Showing Comparison between Control group and Case group (inflammatory biomarker and oxidative stress level Features of the Groups)

**Figure 2:**
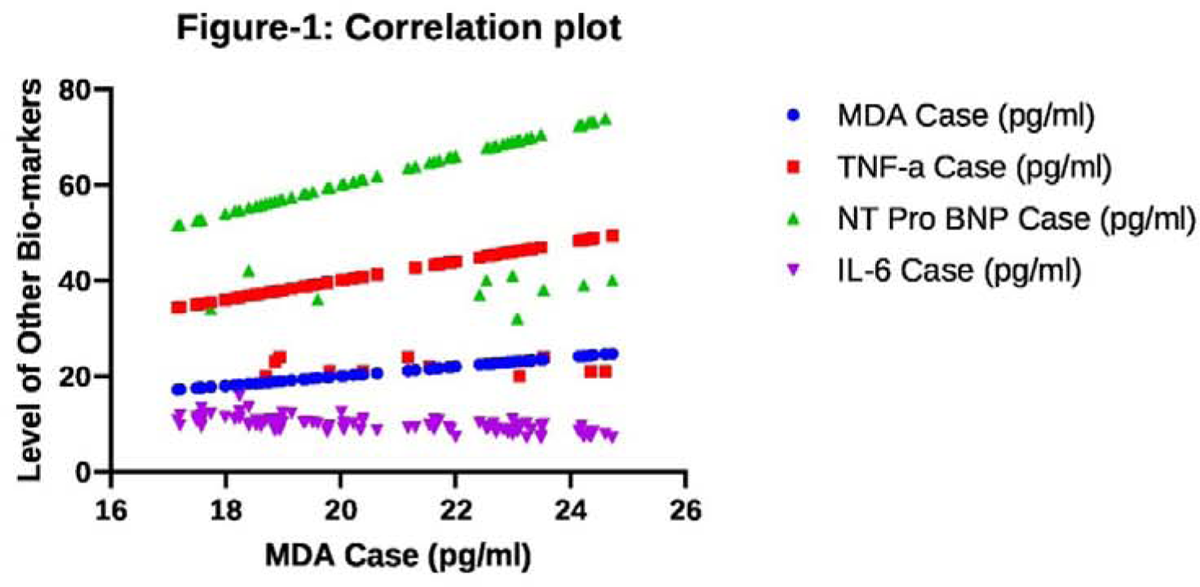
Correlation plot between MDA and Various inflammatory biomarker

**Figure 3:**
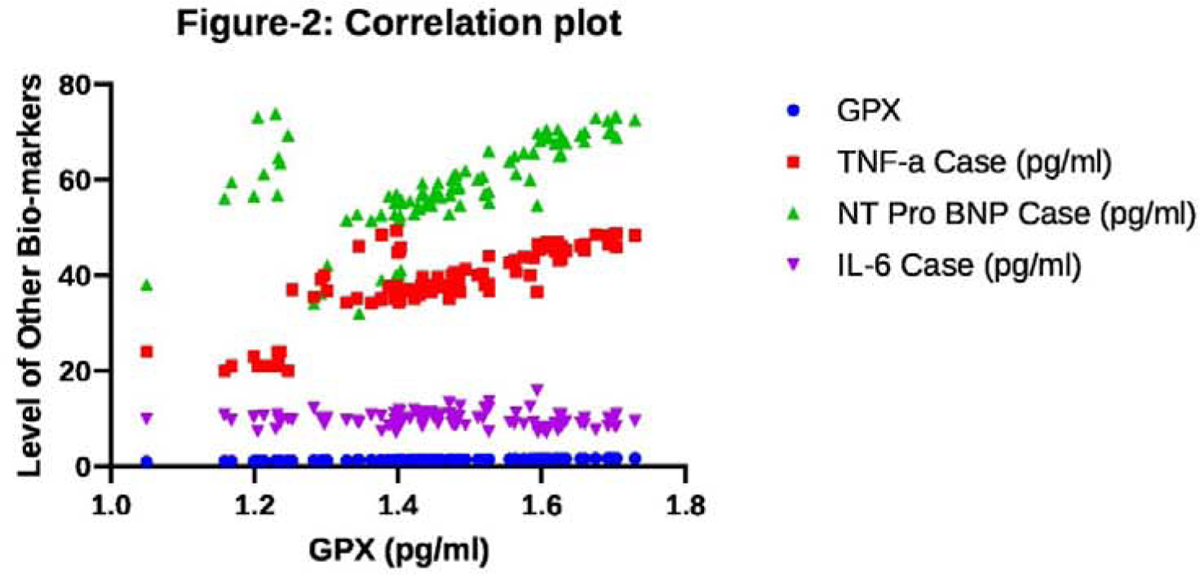
Correlatlon plot between GPX and Various Inflammatory blomarker

**Table 1.** Comparison between Control group and Case group (inflammatory biomarker and oxidative stress level Features of the Groups)

| Variables | Number<br>of values | Minimum | Maximum | *Range* | Mean | Std.<br>Deviation | Std. Error of<br>Mean |
| --- | --- | --- | --- | --- | --- | --- | --- |

|  |  |  |  |  |  |  |  |
| --- | --- | --- | --- | --- | --- | --- | --- |
| MDA Control (pg/ml) | 100 | 0.2795 | 0.5179 | 0.2384 | 0.4100 | 0.07015 | 0.007015 |
| MDA Case (pg/ml) | 100 | 17.16 | 24.73 | 7.569 | 20.58 | 2.308 | 0.2308 |
| CRP Control (pg/ml) | 100 | 2.783 | 3.924 | 1.142 | 3.338 | 0.3168 | 0.03168 |
| CRP Case (pg/ml) | 100 | 21.79 | 31.40 | 9.608 | 26.17 | 2.915 | 0.2915 |
| GPX Control (pg/ml) | 100 | 0.2795 | 0.5179 | 0.2384 | 0.4100 | 0.07015 | 0.007015 |
| GPX Case (pg/ml) | 100 | 1.050 | 1.730 | 0.6799 | 1.466 | 0.1474 | 0.01474 |
| IL-6 Control (pg/ml) | 100 | 1.923 | 2.594 | 0.6716 | 2.236 | 0.1323 | 0.01323 |
| IL-6 Case (pg/ml) | 100 | 7.093 | 15.90 | 8.802 | 9.898 | 1.519 | 1.519 |
| NT Pro BNP Control (pg/ml) | 100 | 0.3590 | 67.00 | 66.64 | 7.902 | 12.16 | 1.216 |
| NT Pro BNP Case (pg/ml) | 100 | 32.00 | 73.83 | 73.83 | 58.55 | 9.997 | 0.9997 |
| TNF-α Control (pg/ml) | 100 | 1.427 | 2.643 | 1.217 | 1.900 | 0.2447 | 0.02447 |
| TNF-α Case (pg/ml) | 100 | 20.00 | 49.47 | 29.47 | 38.86 | 7.422 | 0.7422 |

**Table 2:**
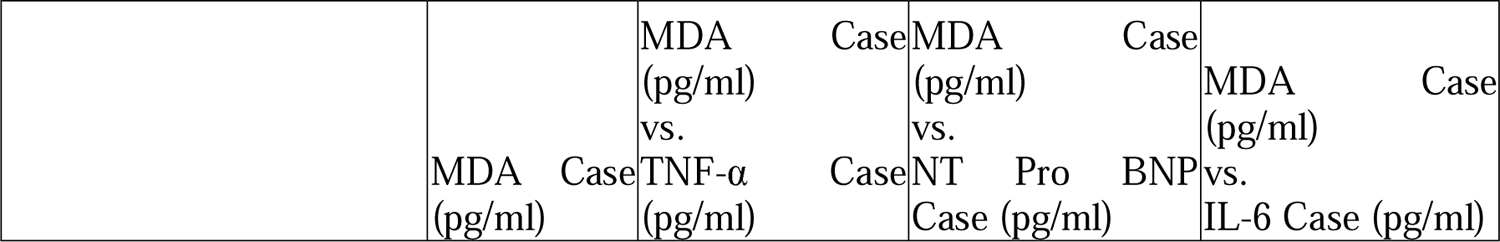
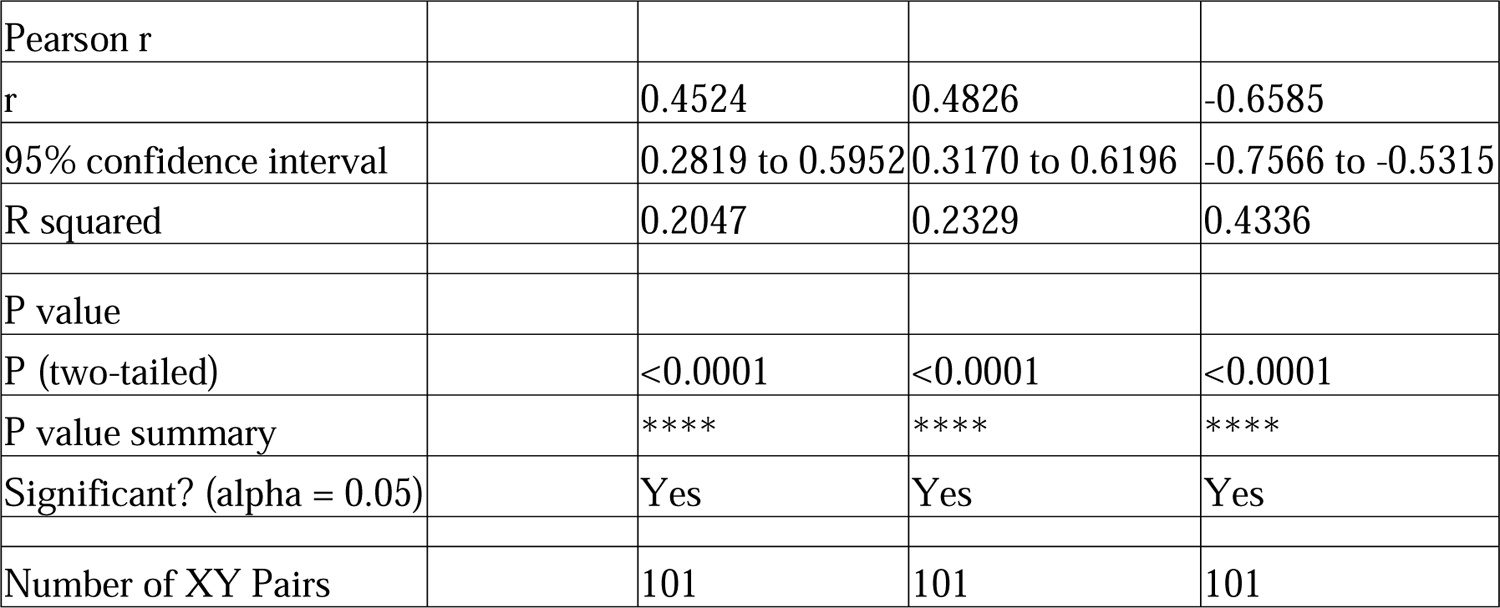
Correlation Analysis between MDA and Various inflammatory biomarker.

**Table 3:**
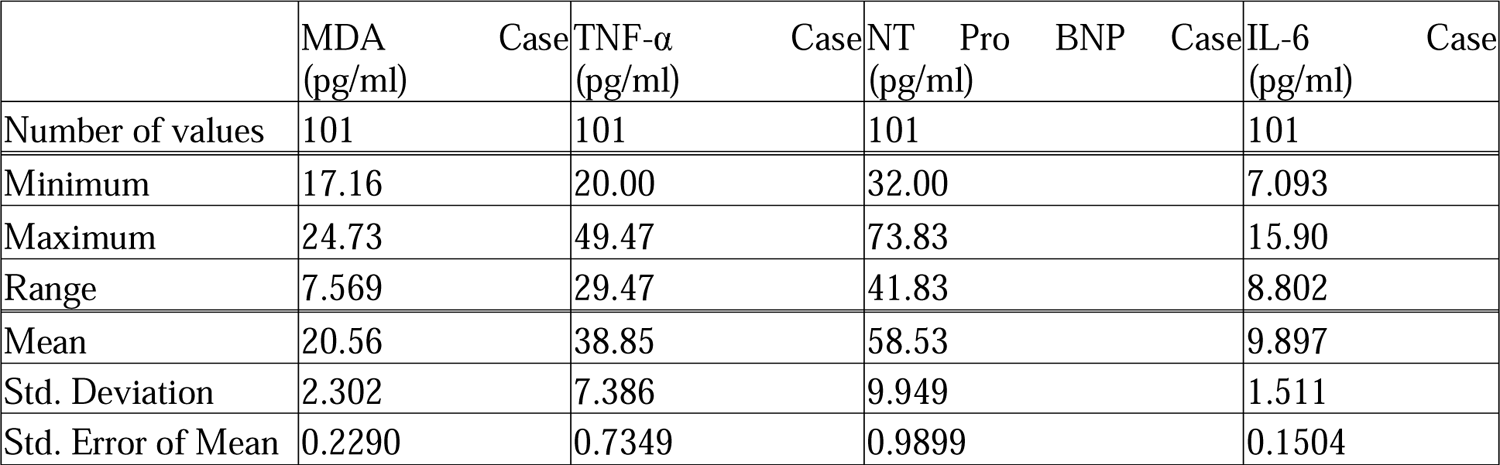
Detail of Correlation of Table-3.

## DISCUSSION

This study uncovers new data on the link between the disease severity marker NT-proBNP and indicators of inflammation and oxidative stress in cardiac patients. Numerous studies shown that local and systemic chronic inflammation is a hallmark of cardiomyopathy.^1,2^ Oxidative stress and chronic inflammation have already been related in human studies to individuals with cardiomyopathy.^1,2,16,20,26^

Numerous inflammatory markers are present in higher amounts in human heart failure patients^1,8,10,12–15,20^ TNF-α, IL-1, and IL-6 are pro-inflammatory cytokines that are especially detrimental to the heart, aggravate hemodynamic abnormalities, and lead to inflammation, tissue loss, and weight loss.^2^ It has been find that individuals with cardiomyopathy have altered levels of inflammatory, pro-, and anti-fibrotic blood mRNA expression.

Our findings revealed that individuals with DCM and other type of cardiomyopathy had considerably greater TNF-α concentrations when compared to healthy controls cases. In a preliminary investigation conducted by Freeman and colleagues^29^, individuals with cardiomyopathy had significantly greater TNF-α concentrations than control cases. Our study employed to assess the levels of cytokine. Our results are corroborated by a research by Freeman and colleagues that did not identify any difference in TNF-α levels between patients with cardiomyopathy and healthy individuals.^30^ TNF-α was not found in any cardiomyopathy patient in the tests of Zois and colleagues or Kim^14^ and colleagues.^31^ Furthermore, there was no significant variation in TNF-α concentration between those with different types of cardiomyopathy.^15^ There are several pro-inflammatory cytokines show high level in the blood of individuals with heart failure this fact based on various studies.^4,25,32^ White blood cell count, WBC, C-reactive protein, and other high-level indicators can all lead to systemic inflammation. Serum CRP concentrations were considerably higher in cardiomyopathy patients compared to controls in our research. CRP levels were observed to be higher in people with different cardiomyopathies and other cardiovascular diseases in previous investigations.^9, 11,12,15,33^ MDA and glutathione peroxidase (GPX) concentrations were considerably greater in cardiomyopathy patients compared to controls in our research. Similar results were reported in another study conducted by in by Freeman and colleagues.^21,22^ Increased MDA concentrations and impaired GPX activity have been documented in human cardiomyopathy^19,38^ patients. Increased MDA concentrations and impaired GPX activity have been documented in human cardiomyopathy patients. We found extremely high correlations in our cardiomyopathy patients, suggesting a connection between oxygen consumption with the inflammatory process. Our results agree with those of Tsutamoto et al., who found that oxidative stress and TNF-α were related in patients with dilated cardiomyopathy. TNF-α promotes ROS production via endothelial mitochondria, NADPH, and the plasma membrane; TNF-α is also involved in the regulation of nitric oxide metabolism.^32^ The significant relationships of TNF-α and NT-proBNP with GPX show that GPX activity rises with increasing inflammation in our cardiomyopathy patients. Conversely, lymphocyte percentages were considerably favorably connected with GPX, whereas lymphocyte count was strongly adversely correlated with MDA. The studies show that inflammation and oxidative stress are linked with cardiomyopathy patients. Our study found a strong relationship between oxidative stress, inflammation, and NT-proBNP in a sample of all cardiac patients. TNF-α, IL-6, WBC, neutrophil and monocyte counts were all shown to be strongly correlated with GPX activity. These findings point to the importance of this antioxidant enzyme in the interaction of inflammatory processes and oxidative stress in cardiac patients. Furthermore, GPX concentration was considerably favorably connected with lymphocyte percentage, but MDA concentration was strongly adversely correlated with lymphocyte count. Furthermore, we identified a strong positive connection between NT-proBNP and GPX, as well as a significant positive correlation between TNF-α and all patients. These findings imply that the severity of the disease is linked to a increase in GPX activity and an increase in inflammation. All of these findings support the hypothesis that inflammation and oxidative stress interact in cardiac patients. Similarly, Rubio and colleagues identified robust relationships between selected inflammatory markers, antioxidants, and echocardiographic variables, suggesting that inflammation and oxidative stress both contribute to cardiomyopathy.^15^In contrast to MDA, we discovered much increased GPX activity in DCM when compared to the control scenario.

This might be because they were significantly older than the DCM patients and control case, or it could be owing to the aetiology of the disorder. Patients with DCM and other cardiomyopathies had significantly greater NT-proBNP concentrations than healthy controls, as would be predicted.

## CONCLUSIONS

In cardiomyopathy patients, but not in normal cases, there is a correlation between inflammatory and oxidative stress indicators. These findings imply intricate cross-talk between the two cellular processes in late-stage cardiomyopathy.

## Data Availability

All data produced in the present work are contained in the manuscript.

## ACKNOWLEDGEMENTS

We are grateful to the Multidisciplinary Research Unit at the Institute of Medical Sciences at Banaras Hindu University for providing us with all the assistance we need for our individual research.

